# Behavioral and psychological symptoms of dementia in different dementia disorders: A large-scale study of 10 000 individuals

**DOI:** 10.1101/2021.06.24.21259454

**Authors:** Emilia Schwertner, Joana B. Pereira, Hong Xu, Juraj Secnik, Bengt Winblad, Maria Eriksdotter, Katarina Nagga, Dorota Religa

## Abstract

**Background:** The majority of individuals with dementia will suffer from psychological and behavioral symptoms of dementia (BPSD). There is limited number of studies investigating BPSD in different types of dementia.

**Objectives:** To characterize BPSD in Alzheimer’s disease (AD), vascular dementia (VaD), mixed (Mixed) dementia, Parkinson’s disease dementia (PDD), dementia with Lewy bodies (DLB), frontotemporal dementia (FTD) and unspecified dementia (UNS) in individuals residing in long-term care facilities.

**Methods:** We included 10,405 individuals with dementia living in long-term care facilities from the Swedish registry for cognitive/dementia disorders (SveDem) and the Swedish BPSD registry. BPSD was assessed with the Neuropsychiatric Inventory - Nursing Home Version (NPI-NH). Multivariate logistic regression models were used to evaluate the associations between dementia diagnoses and different BPSDs.

**Results:** The most common symptoms were aberrant motor behavior, agitation and irritability. Compared to AD, we found a lower risk of delusions (in FTD, unspecified dementia), hallucinations (in FTD), agitation (VaD, in PDD, unspecified dementia), elation/euphoria (in DLB), anxiety (Mixed, VaD, unspecified dementia), disinhibition (in PDD); irritability (in DLB, FTD, unspecified dementia), aberrant motor behavior (in Mixed, VaD, unspecified dementia), sleep and night-time behavior changes (in unspecified dementia). Higher risk of delusions (in DLB), hallucinations (in DLB, PDD), apathy (VaD, FTD), disinhibition (in FTD) and appetite and eating abnormalities (in FTD) were also found in comparison to AD.

**Conclusions:** BPSD was common in all types of dementia, with the most common symptoms being aberrant motor behavior, agitation, and irritability. Additionally, presentation of BPSD differ in different dementia types.

## Introduction

Dementia is a complex syndrome associated with cognitive, behavioral, and psychological symptoms. The current diagnostic system emphasizes cognitive impairment and progressive cognitive decline as the key features of dementia [1]. However, cognitive impairment does not solely explain functional disability or poor quality of life in people with dementia. Instead, recent studies have shown that approximately 90% of individuals with dementia will suffer from additional psychological or behavioral symptoms and that these symptoms are important factors contributing to functional impairment and increased caregiver burden [2–5]. Behavioral and psychological symptoms of dementia (BPSD) (also referred to as neuropsychiatric symptoms of dementia) include changes in behavior, perception, content of thoughts as well as mood disorders [5]. These symptoms are commonly found across different dementia disorders such as Alzheimer’s disease (AD), vascular dementia (VaD), dementia with Lewy bodies (DLB), Parkinson’s disease dementia (PDD) or frontotemporal dementia (FTD). However, studies assessing the prevalence of BPSD in different dementias have reported inconclusive findings. These conflicting results may be attributed to differences in study settings, population demographics, small sample sizes, evaluation methods or the severity of cognitive impairment. Recently, there has been an increasing effort to place BPSD in a more central position in dementia [6,7]. Although studies have reported the presence of these symptoms in AD, VaD, DLB, PDD and FTD, these studies are limited by the lack of comparison of BPSD symptoms between different dementia disorders. Additionally, most of the previous studies were conducted in outpatient populations. Presentation of BPSD can differ between community-dwelling individuals and residents of long-term care facilities. For instance, in AD, the study setting has been previously shown to affect the prevalence of irritability, aberrant motor behaviour, euphoria and appetite disorders [8]. These symptoms might be difficult to manage in a home setting and therefore they significantly contribute to the decision of moving a patient to a long-term care facility [9,10]. The aim of this study was to provide detailed and comprehensive description of presence of BPSD across different dementia diagnoses through a merge of two large Swedish quality registries, in individuals residing in long-term care facilities. The novelty of our work resides on the large number of participants and comparison of BPSD between seven types of dementias, which has not been described to this date.

## Materials and Methods

**The Swedish registry for cognitive/dementia disorders (SveDem)** was established in 2007 and focuses on improving the quality of care of persons with dementia. Individuals are registered at the time of the dementia diagnosis and during follow-up annually in specialist units, primary care centers or in long-term care facilities. Data on type of dementia, age, sex and Mini–Mental State Examination (MMSE) scores at the time of the dementia diagnosis, medication, and living arrangements are collected. Eight types of dementia diagnosis are registered: AD, Mixed, VaD, DLB, PDD, FTD, unspecified dementia, and other dementia types (including various dementia disorders e.g. corticobasal degeneration and alcohol-related dementia). For a more detailed description of SveDem please see Religa et al [11].

**The Swedish BPSD registry** was established in 2010 to improve the quality of care of persons with dementia with BPSD and to achieve a national standard of care throughout Sweden. It includes information about BPSD, current medical treatment and a checklist for possible causes of BPSD [13]. In the registry, assessment of BPSD is carried out using the Neuropsychiatric Inventory-Nursing Home Version (NPI-NH)[14], which includes twelve categories of neuropsychiatric symptoms: delusions, hallucinations, agitation/aggression, depression/depression, apathy, elation/euphoria, anxiety, disinhibition, irritability, aberrant motor behavior, sleep and night-time behavior changes, appetite and eating abnormalities. The NPI-NH is based on the information provided by a caregiver familiar with the behavior of an individual. First, the frequency of symptoms is scored on a four-point scale from 1 to 4 (1: occasionally, 2: less than once a week, 3: very frequently, 4: more than once a day). Second, the severity of the behavior is rated on a three-point scale: mild, moderate, or severe. By multiplying the frequency and severity scores, the NPI-NH yields a rating for each category between 1 and 12. The total NPI score is calculated by adding up the scores across all categories, which can yield a maximum of 144 points. Domain scores of 4 or more are treated as indicative of clinical significance [15]. Information about medical treatment is collected at the time of BPSD assessment. Specifically, the Anatomical Therapeutic Chemical (ATC) codes for the following medications were recorded: analgesics, antipsychotics (ATC code: N05A), anxiolytics (N05B), hypnotics (N05C), antidepressants (N06AA), acetylcholinesterase inhibitors (AChEI) (N06DA), N-Methyl-D-aspartate (NMDA) receptor antagonist (ATC codes: N06DX). Study has ethical approval (2015/2291-31/5).

### Study sample size

In 2016, there were 44 482 persons registered in the BPSD registry and 68 515 in SveDem. After merging the two registries, 13 413 individuals with different dementia diagnosis and assessed for BPSD-symptoms were identified. Age was recorded at the time of dementia diagnosis and at the time of registration in the BPSD-registry. After excluding individuals with incomplete records (n=256), duplicates (n=111), diagnoses prior to 2007 (n=81) and after 2017 (n=1), there were 12 964 individuals. For the purposes of the current study, we additionally excluded persons with other types of dementia (n=329), individuals who were included in the BPSD registry before being registered in SveDem (n=498) and individuals living in community at the time of BPSD assessment (n=1298) or no information about their residency at the time of BPSD was available (n=434). As a result, our final study sample consisted of 10 405 individuals (Supplementary Figure 1).

### Statistical analyses

Continuous variables were reported as mean ± standard deviation (SD), and categorical variables as counts and proportions. Baseline variables from SveDem and BPSD registries were compared across different dementias using Pearson chi-square for proportions and ANOVA for continuous variables. We had a small fraction of missing data ranging from 1.4 in the variable Living arrangement at the time of dementia diagnosis to 8.3 in MMSE at the time of dementia diagnosis (Supplementary Table 1).

Odds ratios (OR) and 95% Confidence Intervals (CIs) of having a clinically significant BPSD (NPI score > 3 in a given domain or category) (versus no symptom) in each diagnosis comparing to AD were estimated with multivariate logistic regression. Then, the analyses were repeated with each diagnosis used as a reference group. All models were adjusted for age, sex, MMSE and time between SveDem and BPSD registries.

## Results

Overall the most common diagnosis in the cohort was AD (34.1%), followed by VaD (16.4%), Mixed (15.6%), DLB (2.3%), PDD (1.7%), and FTD (1.9%). Further, there were 28.5% individuals with unspecified dementia. Mean age at the time of dementia diagnosis was 80.2 ± 7.9 years and 64.8% were women. At the time of dementia diagnosis, 7.3% were being treated with antipsychotics, 30.5% with antidepressants, 18.9% with hypnotics and 13.7% with anxiolytics. In AD, 69% individuals were treated with AChEI, 11.7% with a NMDA antagonist and 2.4% were receiving both NMDA and AChEI. At the time of the diagnosis, 45.6% of all individuals were living alone, 39.8% with somebody else in their households and 14.7% in long-term care facilities. On average, individuals were registered in the BPSD registry after 1013.5 ± 660.2 days from registration in SveDem.

When registered in the BPSD registry, the mean age was 82.4±7.6 years, 16.9% of individuals were being treated with antipsychotics, 44.5% with antidepressants, 18.9% with hypnotics and 19.7% with anxiolytics. In AD, 38.4% were treated with AChEI, 30% with NMDA antagonist and 12.5% were receiving both.

At least one clinically significant symptom (NPI score > 3 in a given domain) was present in 75.5% of individuals and any symptom (NPI score > 0 in a given domain) was present in 92.3% individuals. The most frequent clinically significant symptoms were aberrant motor behavior (38.4%), agitation (33.4%) and irritability (29.7%) (Table 3). Mean of total NPI score was 23.2 ±21.9 points. The highest mean total NPI score was found in FTD (26.3 ± 22.5), followed by DLB (24.9 ± 23.1), AD (24.8 ±2 2.5), PDD (21.7 ± 16.8), Mixed (22.5 ± 22), VaD (22.5 ± 22) and unspecified dementia (21.7 ± 21.2). The characteristics of the sample are provided in Tables 1 and 2.

**TABLE 1.** BASIC CHARACTERISTICS OF THE STUDY POPULATION AT THE TIME OF REGISTRATION IN SVEDEM.

|  | **Total** | **AD**<br>3548 (34.1) | **VaD**<br>1708 (16.4) | **Mixed**<br>1621 (15.6) | **DLB**<br>236 (2.3) | **PDD**<br>122 (1.7) | **FTD**<br>200 (1.9) | **Unspecified**<br>2970 (28.5) | **p-value** |
| --- | --- | --- | --- | --- | --- | --- | --- | --- | --- |
| **Female** | 6739 (64.8) | 2443 (68.9) | 1015 (59.4) | 1066 (65.8) | 118 (50) | 47 (38.5) | 111 (55.5) | 1939 (65.3) | <0.001 |
| **Age, years** | 80.2 (7.9) | 78.3 (8.3) | 81.5 (7.2) | 81.3 (6.7) | 77.6 (6.9) | 75.9 (7) | 69.8 (9) | 82.2 (7.1) | <0.001 |
| **MMSE, score** | 20.1 (5.1) | 20.4 (5.2) | 20.1 (4.9) | 20.1 (5) | 21.3 (4.9) | 20.8 (4.3) | 22.1 (6.1) | 19.4 (5.2) | <0.001 |
| **Time, days to registration in the BPSD-registry** | 1013.5 (660.2) | 1169.6 (682.8) | 895.2 (622.4) | 997.3 (673.4) | 912.3 (611.8) | 951.2 (575) | 929.9 (585.9) | 920.2 (621.9) | <0.001 |
| **Living arrangement at the time of dementia diagnosis** |  |  |  |  |  |  |  |  |  |
| **Alone** | 4674 (45.6) | 1587 (45.2) | 779 (46.1) | 786 (48.8) | 78 (33.2) | 29 (23.8) | 65 (32.8) | 1350 (46.6) |  |
| **With somebody** | 4080 (39.8) | 1590 (45.3) | 577 (34.2) | 644 (40) | 126 (53.6) | 72 (59) | 109 (55.1) | 962 (33.2) | <0.001 |
| **Long-term care facilities** | 1504 (14.7) | 331 (9.4) | 333 (19.7) | 180 (11.2) | 31 (13.2) | 21 (17.2) | 24 (12.1) | 584 (20.2) |  |
| **Antipsychotics** | 717 (7.3) | 186 (5.5) | 114 (7) | 109 (6.9) | 30 (12.9) | 23 (20.2) | 25 (12.7) | 230 (8.6) | <0.001 |
| **Anxiolytics** | 1340 (13.7) | 349 (10.3) | 270 (16.6) | 162 (10.3) | 26 (11.2) | 16 (13.9) | 25 (12.7) | 492 (18.4) | <0.001 |
| **Hypnotics** | 1849 (18.9) | 546 (16.2) | 362 (22.3) | 284 (18.1) | 52 (22.3) | 20 (17.5) | 31 (15.7) | 554 (20.7) | <0.001 |
| **Antidepressants** | 3006 (30.5) | 1001 (29.5) | 590 (36.1) | 414 (26.2) | 81 (34.8) | 45 (39.1) | 68 (34.2) | 807 (29.9) | <0.001 |
| **AChEI** | 4416 (44.3) | 2380 (69) | 151 (9.2) | 789 (49.6) | 174 (74) | 65 (53.7) | 10 (5) | 847 (30.9) | <0.001 |
| **NMDA antagonist** | 1068 (10.8) | 397 (11.7) | 92 (5.7) | 321 (20.2) | 41 (17.7) | 24 (20.5) | 14 (7) | 179 (6.6) | <0.001 |
| **NMDA antagonist + AChEIs** | 151 (1.5) | 81 (2.4) | 4 (0.2) | 20 (1.3) | 15 (6.5) | 9 (7.7) | 1 (0.5) | 21 (0.8) | <0.001 |

Abbreviations: AD - Alzheimer Disease; Mixed - Mixed Dementia; VaD – Vascular dementia; DLB – Dementia with Lewy Bodies; PDD- Parkinson’s disease dementia; FTD – Frontotemporal dementia; MMSE - Mini-Mental State Examination; Living arrangement - at the time of dementia diagnoses; NMDA - N-Methyl-D-aspartate, AChEIs - Acetylcholineesterase inhibitors. Data are presented as mean (SD) or n (%).

**TABLE 2.** BASIC CHARACTERISTICS OF THE STUDY POPULATION AT THE TIME OF REGISTRATION IN THE BPSD REGISTRY.

|  | **Total** | **AD** | **VaD** | **Mixed** | **DLB** | **PDD** | **FTD** | **Unspecified** | **p-value** |
| --- | --- | --- | --- | --- | --- | --- | --- | --- | --- |
| **Age** | 82.4 (7.6) | 80.9 (8) | 83.4 (7) | 83.5 (6.4) | 79.6 (6.6) | 78 (6.9) | 71.8 (8.9) | 84.1 (7) | <0.001 |
| **Analgesics** | 4024 (38.7) | 1297 (36.6) | 713 (41.7) | 636 (39.2) | 89 (37.7) | 47 (38.5) | 58 (29) | 1184 (39.9) | 0.006 |
| **Antipsychotics** | 1757 (16.9) | 615 (17.3) | 279 (16.3) | 242 (14.9) | 44 (18.6) | 33 (27) | 53 (26.5) | 491 (16.5) | <0.001 |
| **Anxiolytics** | 2049 (19.7) | 723 (20.4) | 336 (19.7) | 301 (18.6) | 37 (15.7) | 14 (11.5) | 30 (15) | 608 (20.5) | 0.031 |
| **Hypnotics** | 1967 (18.9) | 672 (18.9) | 346 (20.3) | 337 (20.8) | 52 (22) | 18 (14.8) | 39 (19.5) | 503 (16.9) | 0.013 |
| **Antidepressants** | 4635 (44.5) | 1618 (45.6) | 808 (47.3) | 707 (43.6) | 103 (43.6) | 60 (49.2) | 78 (39) | 1261 (42.5) | 0.012 |
| **AChEI** | 2773 (26.7) | 1361 (38.4) | 116 (6.8) | 519 (32) | 126 (53.4) | 44 (36.1) | 10 (5) | 597 (20.1) | <0.001 |
| **NMDA antagonist** | 2366 (22.7) | 1063 (30) | 186 (10.9) | 496 (30.6) | 92 (39) | 33 (27) | 22 (11) | 474 (16) | <0.001 |
| **NMDA + AChEI** | 808 (7.8) | 444 (12.5) | 16 (0.937) | 150 (9.25) | 58 (24.6) | 15 (12.3) | 4 (2) | 121 (4.07) | <0.001 |
| **Total NPI** | 23.2 (21.9) | 24.8 (22.5) | 22.5 (21.6) | 22.5 (22) | 24.9 (23.1) | 21.7 (16.8) | 26.3 (22.5) | 21.7 (21.2) | <0.001 |
| **Any symptom (NPI >0)** | 9430 (92.3) | 3228 (92.9) | 1529 (91.1) | 1466 (92.2) | 220 (94.4) | 115 (95) | 186 (94.9) | 2686 (91.8) | 0.093 |
| **Clinically significant symptom (NPI >3)** | 7710 (75.5) | 2630 (75.8) | 1249 (74.4) | 1183 (74.4) | 187 (80.6) | 105 (86.8) | 136 (69.4) | 2220 (76) | 0.006 |

Abbreviations: AD - Alzheimer Disease. Mixed - Mixed Dementia, VaD – Vascular dementia, DLB – Dementia with Lewy Bodies, PDD - Parkinson’s disease dementia, FTD – Frontotemporal dementia, NMDA – N-Methyl-D-aspartate, NPI – Neuropsychiatric Inventory. Data are presented as mean (SD) or n (%).

**TABLE 3.**
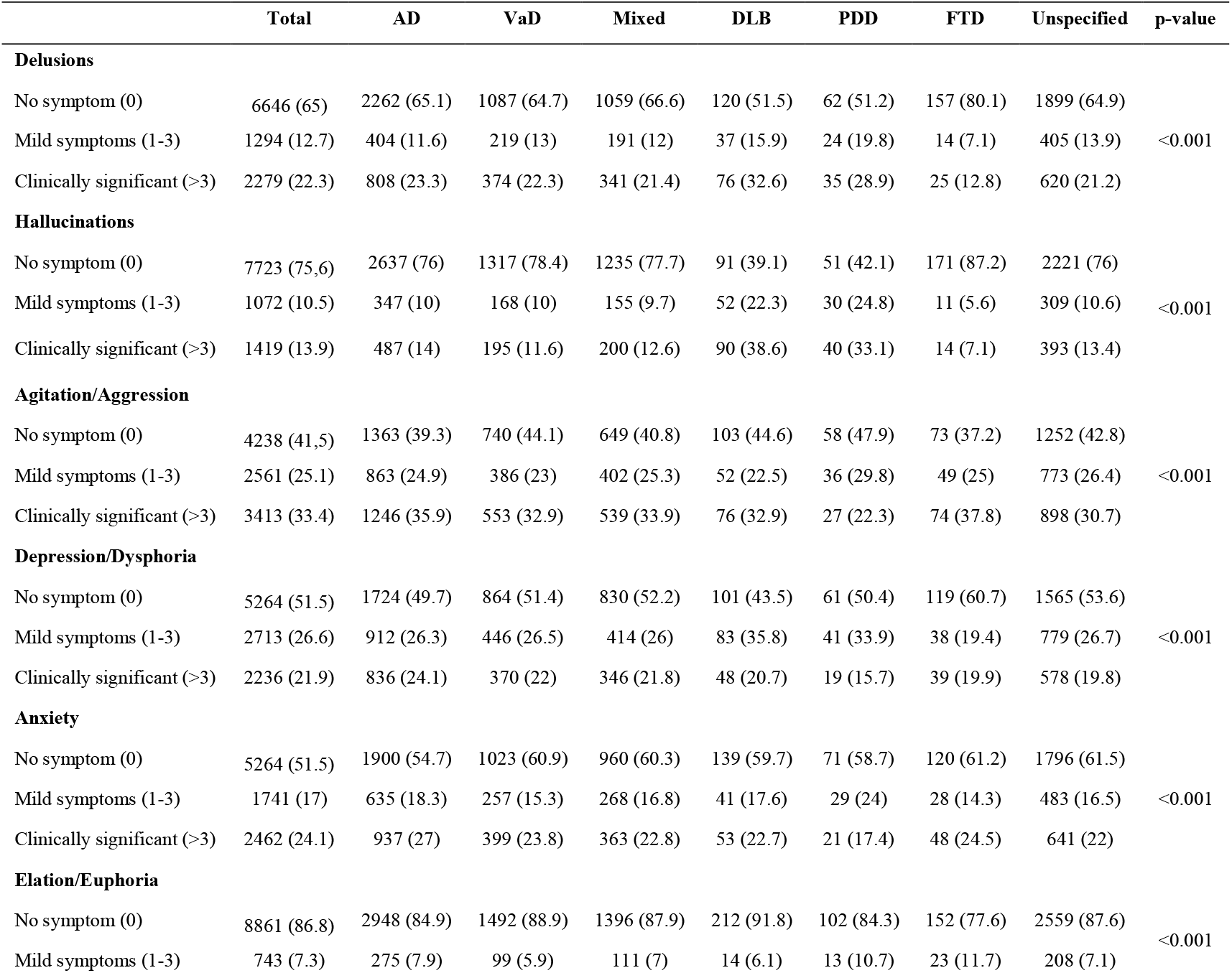
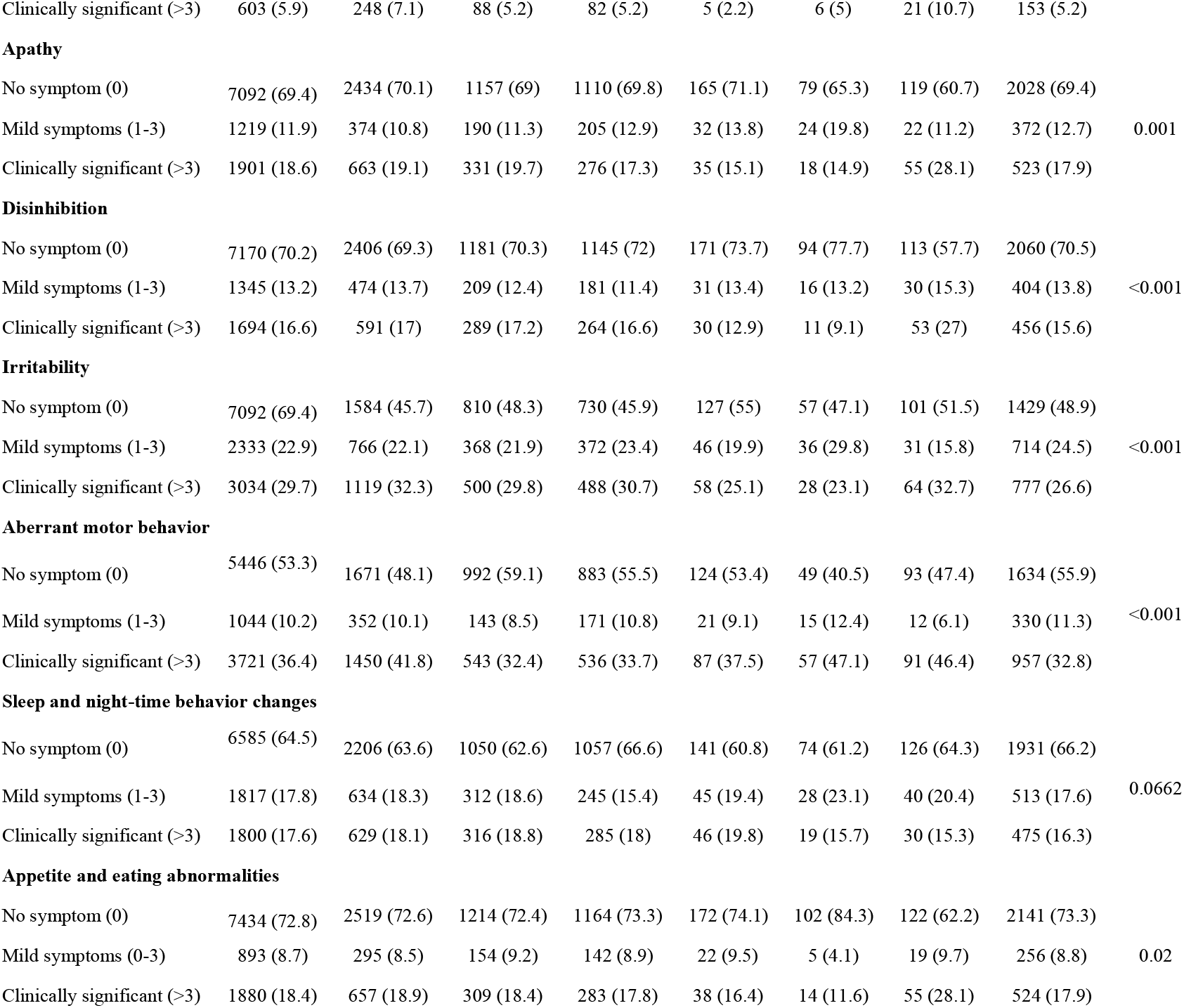
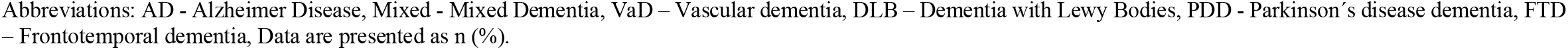
FREQUENCY OF BPSD ACROSS DIFFERENT DEMENTIA DIAGNOSES AND LIVING ARRANGEMENTS.

|  | Total | AD | VaD | Mixed | DLB | PDD | FTD | Unspecified | p-value |
| --- | --- | --- | --- | --- | --- | --- | --- | --- | --- |
| **Delusions** |  |  |  |  |  |  |  |  |  |
| No symptom (0) | 6646 (65) | 2262 (65.1) | 1087 (64.7) | 1059 (66.6) | 120 (51.5) | 62 (51.2) | 157 (80.1) | 1899 (64.9) |  |
| Mild symptoms (1-3) | 1294 (12.7) | 404 (11.6) | 219 (13) | 191 (12) | 37 (15.9) | 24 (19.8) | 14 (7.1) | 405 (13.9) | <0.001 |
| Clinically significant (>3) | 2279 (22.3) | 808 (23.3) | 374 (22.3) | 341 (21.4) | 76 (32.6) | 35 (28.9) | 25 (12.8) | 620 (21.2) |  |
| **Hallucinations** |  |  |  |  |  |  |  |  |  |
| No symptom (0) | 7723 (75.6) | 2637 (76) | 1317 (78.4) | 1235 (77.7) | 91 (39.1) | 51 (42.1) | 171 (87.2) | 2221 (76) |  |
| Mild symptoms (1-3) | 1072 (10.5) | 347 (10) | 168 (10) | 155 (9.7) | 52 (22.3) | 30 (24.8) | 11 (5.6) | 309 (10.6) | <0.001 |
| Clinically significant (>3) | 1419 (13.9) | 487 (14) | 195 (11.6) | 200 (12.6) | 90 (38.6) | 40 (33.1) | 14 (7.1) | 393 (13.4) |  |
| **Agitation/Aggression** |  |  |  |  |  |  |  |  |  |
| No symptom (0) | 4238 (41.5) | 1363 (39.3) | 740 (44.1) | 649 (40.8) | 103 (44.6) | 58 (47.9) | 73 (37.2) | 1252 (42.8) |  |
| Mild symptoms (1-3) | 2561 (25.1) | 863 (24.9) | 386 (23) | 402 (25.3) | 52 (22.5) | 36 (29.8) | 49 (25) | 773 (26.4) | <0.001 |
| Clinically significant (>3) | 3413 (33.4) | 1246 (35.9) | 553 (32.9) | 539 (33.9) | 76 (32.9) | 27 (22.3) | 74 (37.8) | 898 (30.7) |  |
| **Depression/Dysphoria** |  |  |  |  |  |  |  |  |  |
| No symptom (0) | 5264 (51.5) | 1724 (49.7) | 864 (51.4) | 830 (52.2) | 101 (43.5) | 61 (50.4) | 119 (60.7) | 1565 (53.6) |  |
| Mild symptoms (1-3) | 2713 (26.6) | 912 (26.3) | 446 (26.5) | 414 (26) | 83 (35.8) | 41 (33.9) | 38 (19.4) | 779 (26.7) | <0.001 |
| Clinically significant (>3) | 2236 (21.9) | 836 (24.1) | 370 (22) | 346 (21.8) | 48 (20.7) | 19 (15.7) | 39 (19.9) | 578 (19.8) |  |
| **Anxiety** |  |  |  |  |  |  |  |  |  |
| No symptom (0) | 5264 (51.5) | 1900 (54.7) | 1023 (60.9) | 960 (60.3) | 139 (59.7) | 71 (58.7) | 120 (61.2) | 1796 (61.5) |  |
| Mild symptoms (1-3) | 1741 (17) | 635 (18.3) | 257 (15.3) | 268 (16.8) | 41 (17.6) | 29 (24) | 28 (14.3) | 483 (16.5) | <0.001 |
| Clinically significant (>3) | 2462 (24.1) | 937 (27) | 399 (23.8) | 363 (22.8) | 53 (22.7) | 21 (17.4) | 48 (24.5) | 641 (22) |  |
| **Elation/Euphoria** |  |  |  |  |  |  |  |  |  |
| No symptom (0) | 8861 (86.8) | 2948 (84.9) | 1492 (88.9) | 1396 (87.9) | 212 (91.8) | 102 (84.3) | 152 (77.6) | 2559 (87.6) |  |
| Mild symptoms (1-3) | 743 (7.3) | 275 (7.9) | 99 (5.9) | 111 (7) | 14 (6.1) | 13 (10.7) | 23 (11.7) | 208 (7.1) | <0.001 |
| Clinically significant (>3) | 603 (5.9) | 248 (7.1) | 88 (5.2) | 82 (5.2) | 5 (2.2) | 6 (5) | 21 (10.7) | 153 (5.2) |  |
| **Apathy** |  |  |  |  |  |  |  |  |  |
| No symptom (0) | 7092 (69.4) | 2434 (70.1) | 1157 (69) | 1110 (69.8) | 165 (71.1) | 79 (65.3) | 119 (60.7) | 2028 (69.4) |  |
| Mild symptoms (1-3) | 1219 (11.9) | 374 (10.8) | 190 (11.3) | 205 (12.9) | 32 (13.8) | 24 (19.8) | 22 (11.2) | 372 (12.7) | 0.001 |
| Clinically significant (>3) | 1901 (18.6) | 663 (19.1) | 331 (19.7) | 276 (17.3) | 35 (15.1) | 18 (14.9) | 55 (28.1) | 523 (17.9) |  |
| **Disinhibition** |  |  |  |  |  |  |  |  |  |
| No symptom (0) | 7170 (70.2) | 2406 (69.3) | 1181 (70.3) | 1145 (72) | 171 (73.7) | 94 (77.7) | 113 (57.7) | 2060 (70.5) |  |
| Mild symptoms (1-3) | 1345 (13.2) | 474 (13.7) | 209 (12.4) | 181 (11.4) | 31 (13.4) | 16 (13.2) | 30 (15.3) | 404 (13.8) | <0.001 |
| Clinically significant (>3) | 1694 (16.6) | 591 (17) | 289 (17.2) | 264 (16.6) | 30 (12.9) | 11 (9.1) | 53 (27) | 456 (15.6) |  |
| **Irritability** |  |  |  |  |  |  |  |  |  |
| No symptom (0) | 7092 (69.4) | 1584 (45.7) | 810 (48.3) | 730 (45.9) | 127 (55) | 57 (47.1) | 101 (51.5) | 1429 (48.9) |  |
| Mild symptoms (1-3) | 2333 (22.9) | 766 (22.1) | 368 (21.9) | 372 (23.4) | 46 (19.9) | 36 (29.8) | 31 (15.8) | 714 (24.5) | <0.001 |
| Clinically significant (>3) | 3034 (29.7) | 1119 (32.3) | 500 (29.8) | 488 (30.7) | 58 (25.1) | 28 (23.1) | 64 (32.7) | 777 (26.6) |  |
| **Aberrant motor behavior** |  |  |  |  |  |  |  |  |  |
| No symptom (0) | 5446 (53.3) | 1671 (48.1) | 992 (59.1) | 883 (55.5) | 124 (53.4) | 49 (40.5) | 93 (47.4) | 1634 (55.9) |  |
| Mild symptoms (1-3) | 1044 (10.2) | 352 (10.1) | 143 (8.5) | 171 (10.8) | 21 (9.1) | 15 (12.4) | 12 (6.1) | 330 (11.3) | <0.001 |
| Clinically significant (>3) | 3721 (36.4) | 1450 (41.8) | 543 (32.4) | 536 (33.7) | 87 (37.5) | 57 (47.1) | 91 (46.4) | 957 (32.8) |  |
| **Sleep and night-time behavior changes** |  |  |  |  |  |  |  |  |  |
| No symptom (0) | 6585 (64.5) | 2206 (63.6) | 1050 (62.6) | 1057 (66.6) | 141 (60.8) | 74 (61.2) | 126 (64.3) | 1931 (66.2) |  |
| Mild symptoms (1-3) | 1817 (17.8) | 634 (18.3) | 312 (18.6) | 245 (15.4) | 45 (19.4) | 28 (23.1) | 40 (20.4) | 513 (17.6) | 0.0662 |
| Clinically significant (>3) | 1800 (17.6) | 629 (18.1) | 316 (18.8) | 285 (18) | 46 (19.8) | 19 (15.7) | 30 (15.3) | 475 (16.3) |  |
| **Appetite and eating abnormalities** |  |  |  |  |  |  |  |  |  |
| No symptom (0) | 7434 (72.8) | 2519 (72.6) | 1214 (72.4) | 1164 (73.3) | 172 (74.1) | 102 (84.3) | 122 (62.2) | 2141 (73.3) |  |
| Mild symptoms (0-3) | 893 (8.7) | 295 (8.5) | 154 (9.2) | 142 (8.9) | 22 (9.5) | 5 (4.1) | 19 (9.7) | 256 (8.8) | 0.02 |
| Clinically significant (>3) | 1880 (18.4) | 657 (18.9) | 309 (18.4) | 283 (17.8) | 38 (16.4) | 14 (11.6) | 55 (28.1) | 524 (17.9) |  |

Abbreviations: AD - Alzheimer Disease, Mixed - Mixed Dementia, VaD – Vascular dementia, DLB – Dementia with Lewy Bodies, PDD - Parkinson's disease dementia, FTD – Frontotemporal dementia, Data are presented as n (%).

### Associations between dementia type and BPSD

The most common clinically significant symptoms in all dementias were aberrant motor behavior, agitation/aggression and irritability, except for DLB and PDD. In DLB, hallucinations, aberrant motor behavior and delusions were the most frequent symptoms, whereas in PDD aberrant motor behavior, hallucinations and delusions were the most frequent (Table 3). The multivariate logistic regression analyses showed that, compared to AD, individuals with VaD had higher risk of apathy but lower risk of agitation/aggression, anxiety, aberrant motor behavior; individuals with Mixed dementia had lower risk of clinically significant anxiety and aberrant motor behavior; individuals with DLB had higher risk of delusions and hallucinations and lower risk of elation/euphoria and irritability; individuals with PDD had higher risk of hallucinations but lower risk of agitation/ aggression and disinhibition, individuals with FTD had higher risk of apathy, disinhibition, appetite and eating abnormalities as well as lower risk delusion, hallucinations, depression/dysphoria; individuals with unspecified dementia had lower risk of delusion, agitation/aggression, depression/dysphoria, anxiety, irritability, aberrant motor behavior, sleep and night-time behavior changes (Table 4).

**TABLE 4.** ODDS RATIOS (OR) AND 95% CONFIDENCE INTERVALS (CI) OF THE ASSOCIATION BETWEEN DEMENTIA TYPE AND BPSD WITH THE REFERENCE TO AD IN INDIVIDIALS RESIDING IN LONG-TERM CARE FACILITY.

|  | VaD |  | Mixed |  | DLB |  | PDD |  | FTD |  | Unspecified |  |
| --- | --- | --- | --- | --- | --- | --- | --- | --- | --- | --- | --- | --- |
|  | OR (95%CI) | p-value | OR (95%CI) | p-value | OR (95%CI) | p-value | OR (95%CI) | p-value | OR (95%CI) | p-value | OR (95%CI) | p-value |
| **Delusions** | 0.95 (0.82-1.11) | 0.542 | 0.91 (0.78-1.06) | 0.234 | 1.71 (1.25-2.33) | 0.001 | 1.35 (0.85-2.1) | 0.185 | 0.44 (0.27-0.68) | <0.001 | 0.87 (0.77-1) | 0.044 |
| **Hallucinations** | 0.83 (0.69-1.01) | 0.066 | 0.94 (0.78-1.13) | 0.54 | 5.94 (4.31-8.2) | <0.001 | 4.47 (2.86-6.95) | <0.001 | 0.5 (0.27-0.85) | 0.016 | 1.01 (0.86-1.18) | 0.93 |
| **Agitation/Aggression** | 0.85 (0.73-0.98) | 0.029 | 1.01 (0.87-1.17) | 0.905 | 0.84 (0.61-1.16) | 0.291 | 0.46 (0.28-0.75) | 0.002 | 0.97 (0.67-1.41) | 0.871 | 0.85 (0.75-0.96) | 0.011 |
| **Depression/Dysphoria** | 0.94 (0.8-1.1) | 0.465 | 0.93 (0.79-1.09) | 0.361 | 1.01 (0.69-1.45) | 0.964 | 0.69 (0.39-1.16) | 0.181 | 0.57 (0.38-0.85) | 0.007 | 0.83 (0.72-0.95) | 0.007 |
| **Elation/Euphoria** | 0.9 (0.69-1.18) | 0.463 | 0.89 (0.68-1.16) | 0.395 | 0.32 (0.11-0.71) | 0.014 | 0.79 (0.3-1.69) | 0.58 | 1.25 (0.72-2.05) | 0.405 | 0.89 (0.7-1.11) | 0.307 |
| **Anxiety** | 0.85 (0.73-0.98) | 0.031 | 0.82 (0.71-0.96) | 0.012 | 0.83 (0.58-1.16) | 0.282 | 0.64 (0.37-1.06) | 0.097 | 0.73 (0.49-1.07) | 0.11 | 0.75 (0.66-0.85) | <0.001 |
| **Apathy** | 1.21 (1.03-1.42) | 0.023 | 1.04 (0.88-1.22) | 0.684 | 0.87 (0.58-1.26) | 0.471 | 0.89 (0.5-1.49) | 0.678 | 1.54 (1.06-2.22) | 0.022 | 1.09 (0.94-1.25) | 0.256 |
| **Disinhibition** | 1.03 (0.87-1.22) | 0.714 | 1.02 (0.86-1.21) | 0.816 | 0.71 (0.46-1.05) | 0.097 | 0.4 (0.19-0.74) | 0.007 | 1.52 (1.03-2.2) | 0.03 | 0.97 (0.83-1.12) | 0.683 |
| **Irritability** | 0.94 (0.81-1.08) | 0.381 | 1.08 (0.93-1.24) | 0.322 | 0.67 (0.47-0.93) | 0.018 | 0.65 (0.4-1.04) | 0.081 | 0.69 (0.48-0.98) | 0.042 | 0.83 (0.73-0.95) | 0.005 |
| **Aberrant motor behavior** | 0.66 (0.58-0.76) | <0.001 | 0.75 (0.66-0.86) | <0.001 | 0.75 (0.55-1) | 0.053 | 1.12 (0.74-1.68) | 0.594 | 0.83 (0.59-1.15) | 0.257 | 0.73 (0.65-0.82) | <0.001 |
| **Sleep and night-time behavior changes** | 0.92 (0.78-1.08) | 0.317 | 0.91 (0.77-1.07) | 0.267 | 0.98 (0.67-1.4) | 0.917 | 0.66 (0.37-1.13) | 0.148 | 0.82 (0.51-1.26) | 0.381 | 0.82 (0.71-0.95) | 0.008 |
| **Appetite and eating abnormalities** | 1.08 (0.92-1.27) | 0.33 | 1.02 (0.87-1.2) | 0.809 | 0.92 (0.62-1.32) | 0.649 | 0.63 (0.34-1.07) | 0.109 | 1.84 (1.27-2.63) | 0.001 | 1 (0.87-1.15) | 0.998 |

Abbreviations: AD - Alzheimer Disease, Mixed - Mixed Dementia, VaD – Vascular dementia, DLB – Dementia with Lewy Bodies, PDD - Parkinson’s disease dementia, FTD – Frontotemporal dementia. Model adjusted for age, sex, MMSE and time between registration to SveDem and BPSD registry.

The OR 95% (CI) and p-values for comparisons between other diagnoses are presented in Supplementary Tables 2a-f. The summary of these differences is shown in Table 5.

**Table 5.**
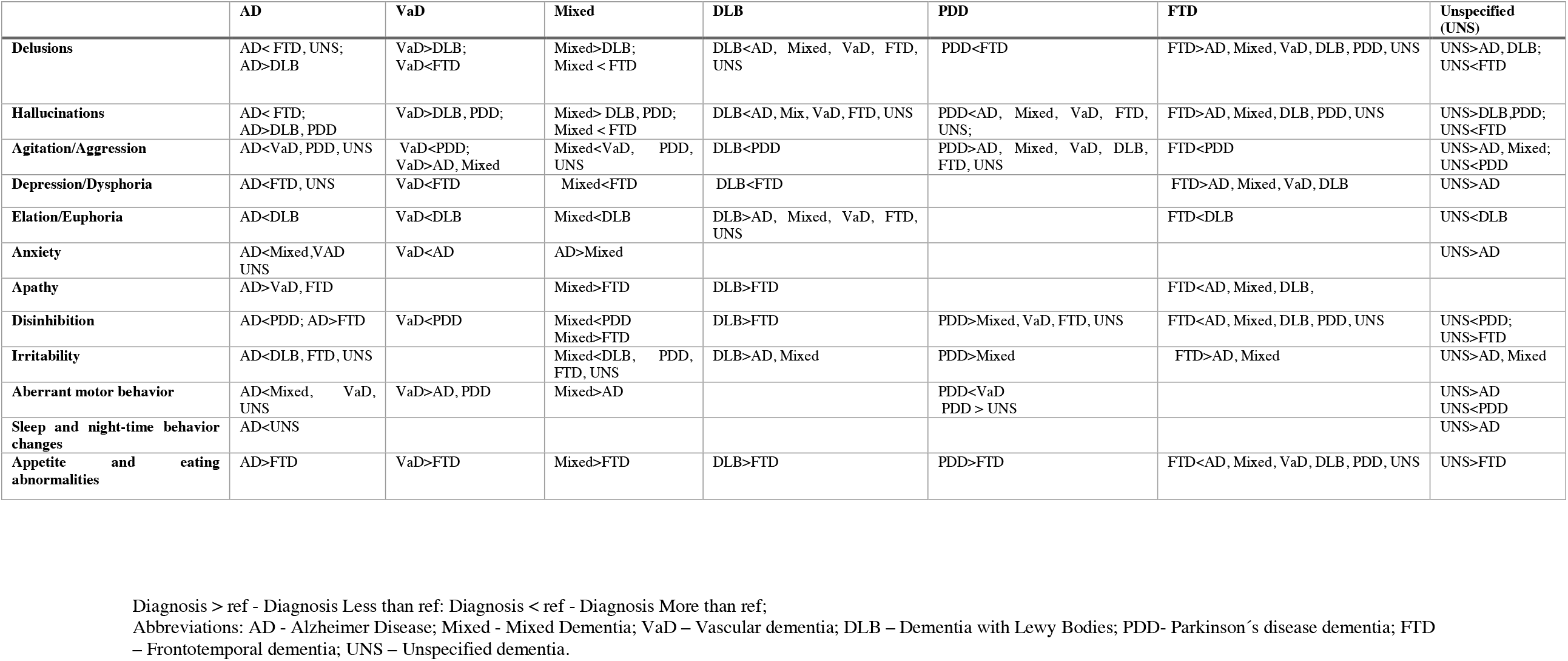
SUMMARY OF THE DIFFERENCES IN BPSD IN INDIVIDIALS RESIDING IN LONG-TERM BASED ON LOGISTIC REGRESSION ANALYSES.

|  | **AD** | **VaD** | **Mixed** | **DLB** | **PDD** | **FTD** | **Unspecified (UNS)** |
| --- | --- | --- | --- | --- | --- | --- | --- |
| **Delusions** | AD< FTD, UNS;<br>AD>DLB | VaD>DLB;<br>VaD<FTD | Mixed>DLB;<br>Mixed < FTD | DLB<AD, Mixed, VaD, FTD, UNS | PDD<FTD | FTD>AD, Mixed, VaD, DLB, PDD, UNS | UNS>AD, DLB;<br>UNS<FTD |
| **Hallucinations** | AD< FTD;<br>AD>DLB, PDD | VaD>DLB, PDD; | Mixed> DLB, PDD;<br>Mixed < FTD | DLB<AD, Mix, VaD, FTD, UNS | PDD<AD, Mixed, VaD, FTD, UNS; | FTD>AD, Mixed, DLB, PDD, UNS | UNS>DLB,PDD;<br>UNS<FTD |
| **Agitation/Aggression** | AD<VaD, PDD, UNS | VaD<PDD;<br>VaD>AD, Mixed | Mixed<VaD, PDD, UNS | DLB<PDD | PDD>AD, Mixed, VaD, DLB, FTD, UNS | FTD<PDD | UNS>AD, Mixed;<br>UNS<PDD |
| **Depression/Dysphoria** | AD<FTD, UNS | VaD<FTD | Mixed<FTD | DLB<FTD |  | FTD>AD, Mixed, VaD, DLB | UNS>AD |
| **Elation/Euphoria** | AD<DLB | VaD<DLB | Mixed<DLB | DLB>AD, Mixed, VaD, FTD, UNS |  | FTD<DLB | UNS<DLB |
| **Anxiety** | AD<Mixed,VAD<br>UNS | VaD<AD | AD>Mixed |  |  |  | UNS>AD |
| **Apathy** | AD>VaD, FTD |  | Mixed>FTD | DLB>FTD |  | FTD<AD, Mixed, DLB, |  |
| **Disinhibition** | AD<PDD; AD>FTD | VaD<PDD | Mixed<PDD<br>Mixed>FTD | DLB>FTD | PDD>Mixed, VaD, FTD, UNS | FTD<AD, Mixed, DLB, PDD, UNS | UNS<PDD;<br>UNS>FTD |
| **Irritability** | AD<DLB, FTD, UNS |  | Mixed<DLB, PDD, FTD, UNS | DLB>AD, Mixed | PDD>Mixed | FTD>AD, Mixed | UNS>AD, Mixed |
| **Aberrant motor behavior** | AD<Mixed, VaD, UNS | VaD>AD, PDD | Mixed>AD |  | PDD<VaD<br>PDD > UNS |  | UNS>AD<br>UNS<PDD |
| **Sleep and night-time behavior changes** | AD<UNS |  |  |  |  |  | UNS>AD |
| **Appetite and eating abnormalities** | AD>FTD | VaD>FTD | Mixed>FTD | DLB>FTD | PDD>FTD | FTD<AD, Mixed, VaD, DLB, PDD, UNS | UNS>FTD |

Diagnosis > ref - Diagnosis Less than ref: Diagnosis < ref - Diagnosis More than ref;
Abbreviations: AD - Alzheimer Disease; Mixed - Mixed Dementia; VaD – Vascular dementia; DLB – Dementia with Lewy Bodies; PDD- Parkinson’s disease dementia; FTD – Frontotemporal dementia; UNS – Unspecified dementia.

## Discussion

In this large-scale study of 10 000 individuals, we observed that BPSD was common across all dementia types, with 75.5% of all individuals exhibiting at least one clinically significant BPSD. Amongst all symptoms, aberrant motor behaviour, agitation and irritability were the most frequent, occurring commonly in all dementias. It has been suggested that brain regions underlying these symptoms might be associated with problems in the inhibition of one’s actions [16]. Moreover, hyperactivity symptoms may be the most important reason of moving to a nursing home and therefore their occurrence may be higher in care facilities[17]. These findings highlight the need of developing strategies for more efficient interventions for these BPSD in long-term facilities.

### Associations between dementia type and BPSD

The main aim of this study was to compare BPSD in different dementia diagnoses. Due to their similar presentation of BPSD, AD, Mixed and VaD are discussed as one group and DLB and PDD as another group.

### Alzheimer disease, Mixed dementia and Vascular dementia

In the present study, the most common clinically significant symptoms in AD, Mixed and VaD were aberrant motor behavior, agitation/aggression and irritability. We found that AD individuals had a higher risk of anxiety, aberrant motor behavior compared with Mixed and VaD as well as agitation in comparison with VaD. Individuals with VaD had higher risk of apathy in comparison to AD. The higher risk of agitation/ aggression in Mixed compared to VaD was the only difference found between these two diagnoses. Although previous studies explored the differences between AD, Mixed and VaD [18–26], their results were not conclusive. Additionally, to our knowledge only three of the previous studies used analyses adjusted for potential confounders [21,23,24] of which two used NPI to assess the symptoms [23,24]. Our study is in line with the work by Caputo et al. (2008)[24] regarding a higher risk of anxiety, agitation and aberrant motor behavior in AD than in VaD. In the study by Fuh et al. (2005)[23] the mean composite scores in sleep disturbance domains in individuals with cortical VaD were higher than those in individuals with AD. Futher, there was a trend for patients with cortical VaD and subcortical VaD to have higher scores tha AD in apathy. However, in our study it was not possible to differentiate between different subtypes of VaD, which could explain why we did not find similar results to previous studies as our VaD patients included individuals with both cortical and subcortical pathology.

### Dementia with Lewy bodies and Parkinson’s disease dementia

Although DLB and PDD share the core characteristics of DLB (hallucinations, cognitive fluctuations, dementia and parkinsonism)[27], previous studies observed a higher mean total NPI score as well as higher frequencies of delusions, hallucinations, agitation, anxiety, irritability, aberrant motor behavior in DLB compared to PDD [28–30]. In our study, apart from a lower risk of agitation/aggression in individuals with PDD, we did not find any other difference between these two diagnoses. In line with previous reports, a higher risk of hallucinations and delusions in DLB compared to other diagnoses was also found in our study [24,31,32]. On the other hand, we found that individuals with DLB had a lower risk of elation/euphoria relative to all other types of dementia, except for PDD. Individuals with PDD had a higher risk of hallucinations than other types of dementia, a higher risk of delusion than FTD and a higher risk of aberrant motor behavior than VaD and unspecified dementia. On the other hand, they also had a lower risk of anxiety and other symptoms from the hyperactivity spectrum (agitation/aggression, disinhibition and irritability). This is partly in line with previous studies reporting a higher rate of agitation, disinhibition, irritability, euphoria in individuals with AD than those with PDD [33]. However, unlike previous studies, more severe aberrant motor behavior in AD[33] and a higher rate of depression in PDD than in AD[34] were not detected in our cohort. Future studies are needed to assess the nature of these discrepancies.

### Frontotemporal dementia

The most frequent symptoms in FTD were aberrant motor behavior, agitation/aggression, irritability and apathy. Furthermore, compared with the other dementia diagnoses, we found a higher risk of apathy (except for VaD, PDD and unspecified dementia), disinhibition (except for VaD), appetite and eating abnormalities in addition to a lower risk of delusions, hallucinations (except for VaD) and depression (except for PDD and unspecified dementia). We also found a higher risk of elation/euphoria in FTD in comparison to DLB. Our results agree with the findings reported by previous studies [25,31,35–38]. There is consistent evidence showing that in individuals with FTD, apathy, disinhibition and appetite and eating abnormalities are frequent symptoms, with abnormal behavior being the core diagnostic feature of FTD [39]. Moreover, in FTD, behavioral changes are one of the core symptoms of the disease, which explains the highest total NPI score found in our study in this group.

### Unspecified dementia

Aberrant motor behavior, agitation/aggression and irritability were the most common clinically significant symptoms in unspecified dementia. Additionally, the lowest total NPI score was found in this group. This was in line with the lower risk of several symptoms in unspecified dementia compared with other types of dementia observed in our cohort. On the other hand, individuals with unspecified dementia had higher risk of elation/euphoria than DLB and disinhibition than PDD. However, these differences were likely driven by the generally low risk of elation/euphoria and disinhibition in these diagnoses. The diagnosis of unspecified dementia is often set in a primary care setting without access to more extensive investigations needed to reveal the etiology of dementia [40]. The unspecified dementia group may therefore include people with various pathologies and thus lack any specific patterns of BPSD.

### General discussion and limitations

Our study was based on the merging of two large quality registries, which enabled a comprehensive large-scale comparison of BPSD between all major subtypes of dementia in a single study. To our knowledge, this is the most comprehensive study on BPSD performed so far. Similar profiles of BPSD in AD, Mixed, VaD and DLB and PDD could be explained by shared pathophysiologic mechanisms [41–43]. Moreover, the presence of symptoms in VaD may vary due to the variability of the location and the extension of ischaemic damage.[44] The VaD cohort in our study was heterogenous and included cases with multi-infarct dementia, strategic infarct dementia as well as subcortical ischemic dementia. There is also a frequent coexistence of Alzheimer’s disease pathology with VaD, DLB and PDD [41–43]. Additionally, potential diagnostic accuracy problems could contribute to these results. For example, some DLB individuals may be misdiagnosed as AD and vice-versa. Moreover, information about the severity of dementia at the time of assessment of BPSD was not available and therefore we used the measurement at the time of dementia diagnosis. The cross-sectional design with a single BPSD measure may obscure differences between different dementia diagnoses. Therefore, a longitudinal design with repeated BPSD assessments should complement the present study. The different sample sizes and their characteristics, study designs and methods to estimate BPSD used in previous studies render the comparisons between our results and previous findings difficult. For instance, in contrast to most previous studies, the majority of individuals in our cohort was older. Furthermore, we cannot rule out that the differences between our and previous studies are due to cultural variation in BPSD [45]. Moreover, we had a fraction of missing data, which was dealt with using the multiple imputation method.

### Conclusions and Implications

In this study, BPSD were frequent in all major types of dementias. The most frequent BPSD were agitation/aggression, aberrant motor behavior and irritability. Additionally, we identified differences in individual BPSD between the dementia diagnoses. Description of BPSD in individuals with dementia may allow for a better understanding of the pathophysiology of the disorder itself. Furthermore, it will also contribute to more personalized care and better quality of individuals with dementia and their caregivers.

## Data Availability

The data underlying this article cannot be shared publicly due to their containing information that could compromise the privacy of research participants.

## Acknowledgements

SveDem is supported financially by the Swedish Associations of Local Authorities and Regions, Gun och Bertil Stohnes Stiftelse, CIMED grant, Alzheimerfonden, Swedish Brain Foundation, and Margaretha af Ugglas’ foundation and Swedish Research Council (Drn 2012-2291 and Drn 2016-02317), and by grants provided by the Stockholm County Council (ALF project).

The authors are grateful to the Swedish Dementia Registry (SveDem, www.svedem.se). We thank all patients, caregivers, reporting units and coordinators in SveDem as well as SveDem steering committee.

**The authors declare no conflicts of interest.**

## Ethics Statement

The authors assert that all procedures contributing to this work comply with the ethical standards of the relevant national and institutional committees on human experimentation and with the Helsinki Declaration of 1975, as revised in 2008. All procedures involving human subjects/patients were approved by the regional ethical review board in Stockholm, Sweden with ethical number: 2015/2291-31/5. All patients receive information about the use of their information and have the right to refuse participation or withdraw their data from the registry at any time.

---

**Supplementary Figure 1.**
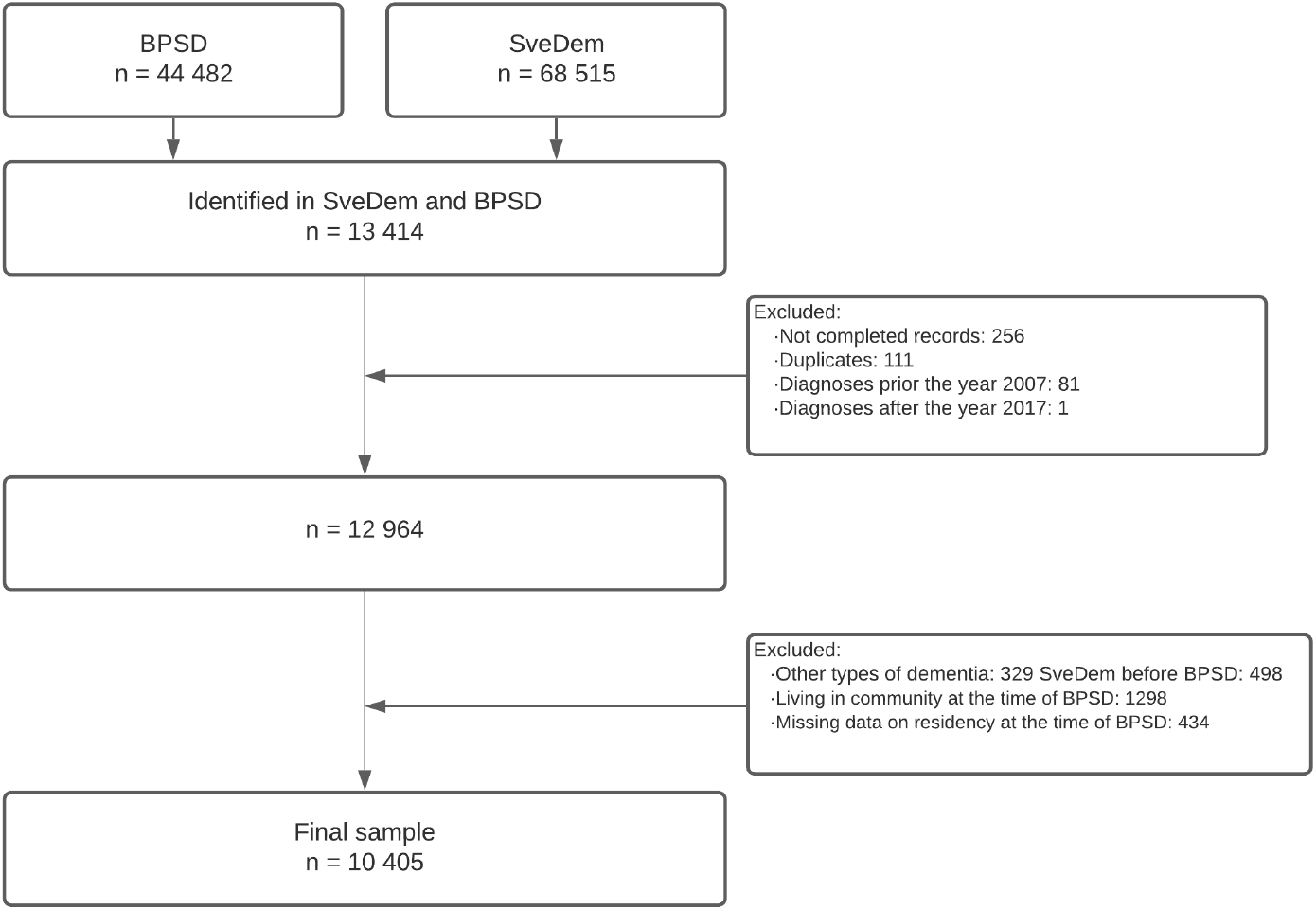
SAMPLE SIZE.

**Supplementary Table 1.** FREQUENCY OF MISSING VALUES IN SVEDEM AND BPSD REGISTRY.

| **Variable** | **n (%)** |
| --- | --- |
| **MMSE at the time of dementia diagnosis** | 1009 (8.3) |
| **Living arrangement at the time of dementia diagnosis** | 169 (1.4) |
| **Sleep and night-time behavior changes** | 245 (2.0) |
| **Irritability** | 241 (2) |
| **Appetite and eating abnormalities** | 241 (2) |
| **Euphoria/Elation** | 240 (2) |
| **Disinhibition** | 237 (2) |
| **Aberrant motor behavior** | 235 (1.9) |
| **Apathy** | 234 (1.9) |
| **Depression/Dysphoria** | 233 (1.9) |
| **Anxiety** | 233 (1.9) |
| **Agitation/Aggression** | 232 (1.9) |
| **Hallucinations** | 231 (1.9) |
| **Delusions** | 225 (1.9) |

MMSE - Mini-Mental State Examination

**Supplementary Table 2a.**
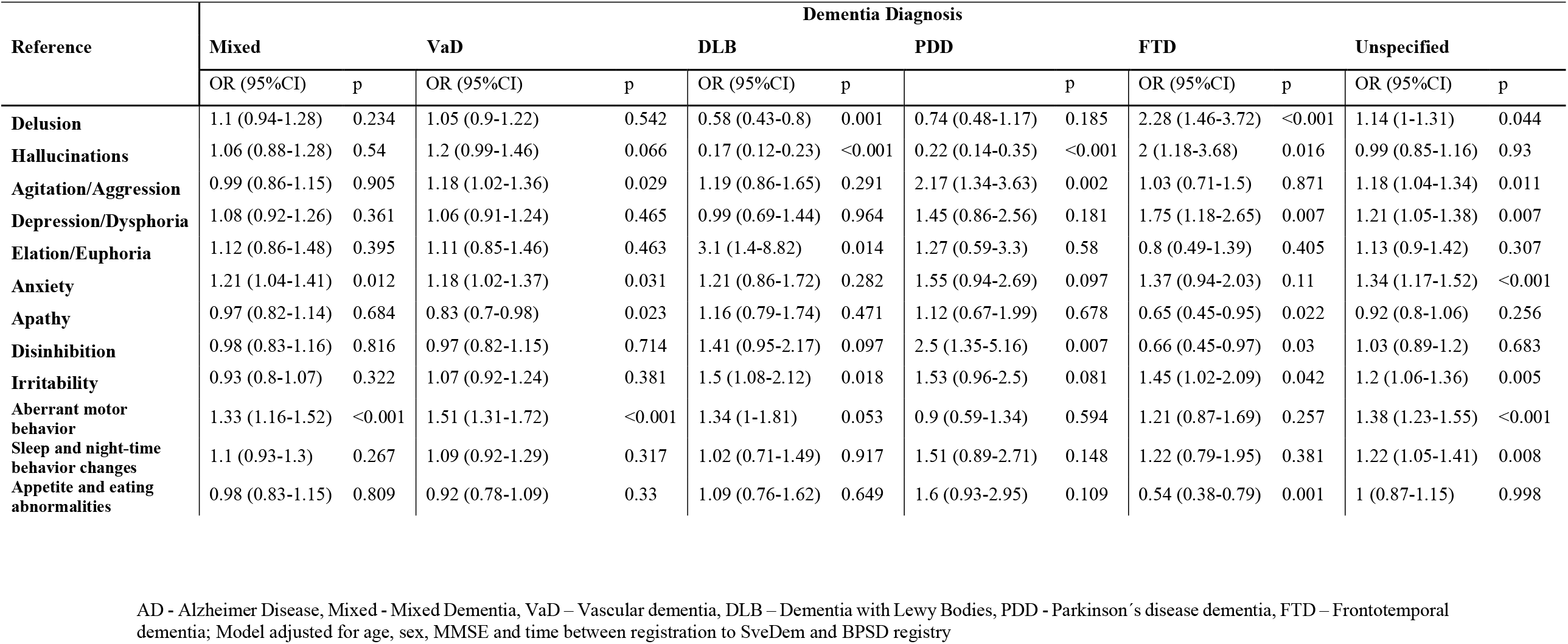
ODDS RATIOS (OR) AND 95% CONFIDENCE INTERVALS (CI) OF THE ASSOCIATION BETWEEN AD AND BPSD COMPARED TO ALL OTHER DIAGNOSES.

**Supplementary Table 2b.**
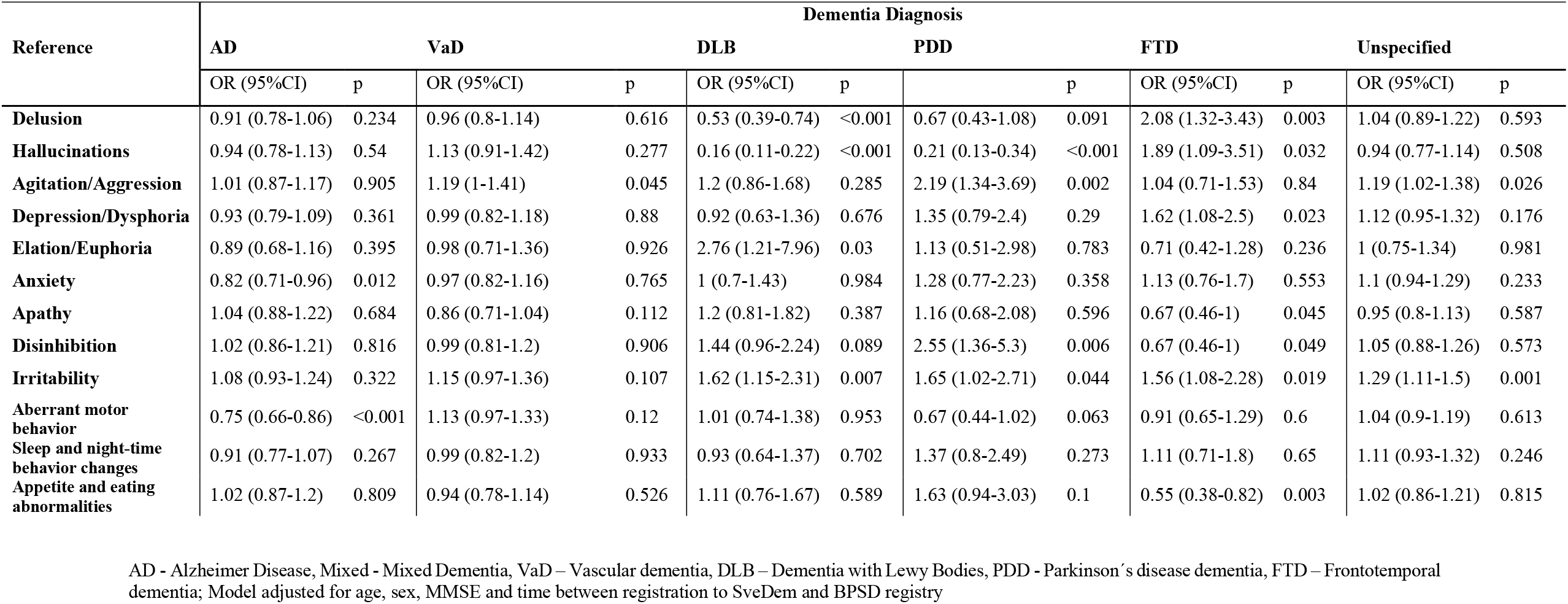
ODDS RATIOS (OR) AND 95% CONFIDENCE INTERVALS (CI) OF THE ASSOCIATION BETWEEN MIXED DEMENTIA AND BPSD COMPARED TO ALL OTHER DIAGNOSES.

**Supplementary Table 2c.**
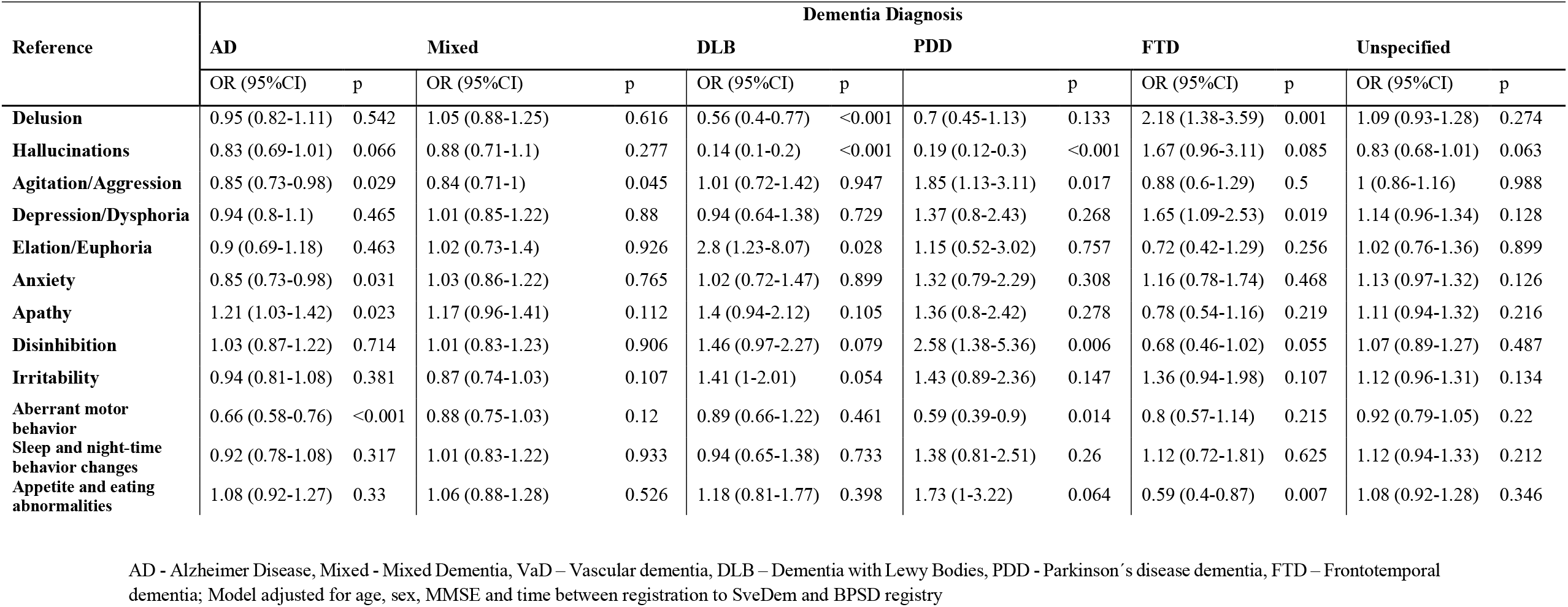
ODDS RATIOS (OR) AND 95% CONFIDENCE INTERVALS (CI) OF THE ASSOCIATION BETWEEN VaD AND BPSD COMPARED TO ALL OTHER DIAGNOSES.

**Supplementary Table 2d.**
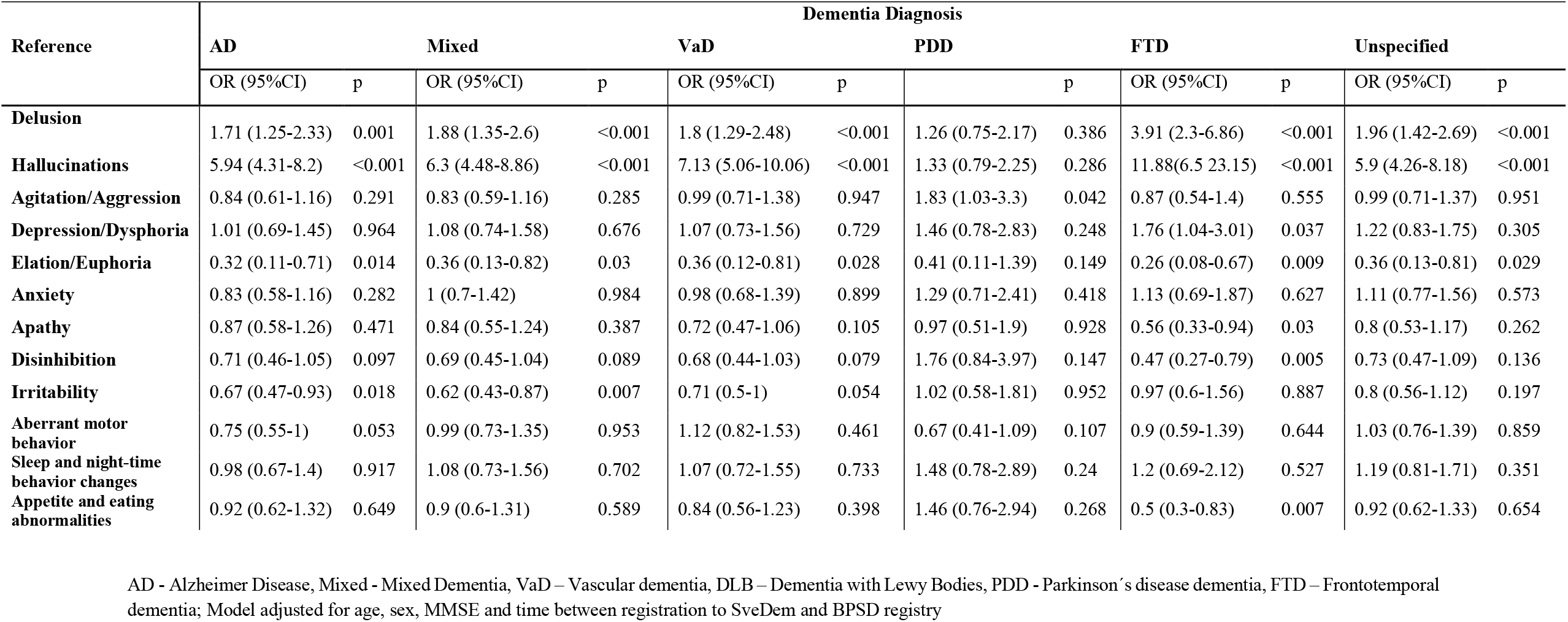
ODDS RATIOS (OR) AND 95% CONFIDENCE INTERVALS (CI) OF THE ASSOCIATION BETWEEN DLB AND BPSD COMPARED TO ALL OTHER DIAGNOSES.

**Supplementary Table 2e.**
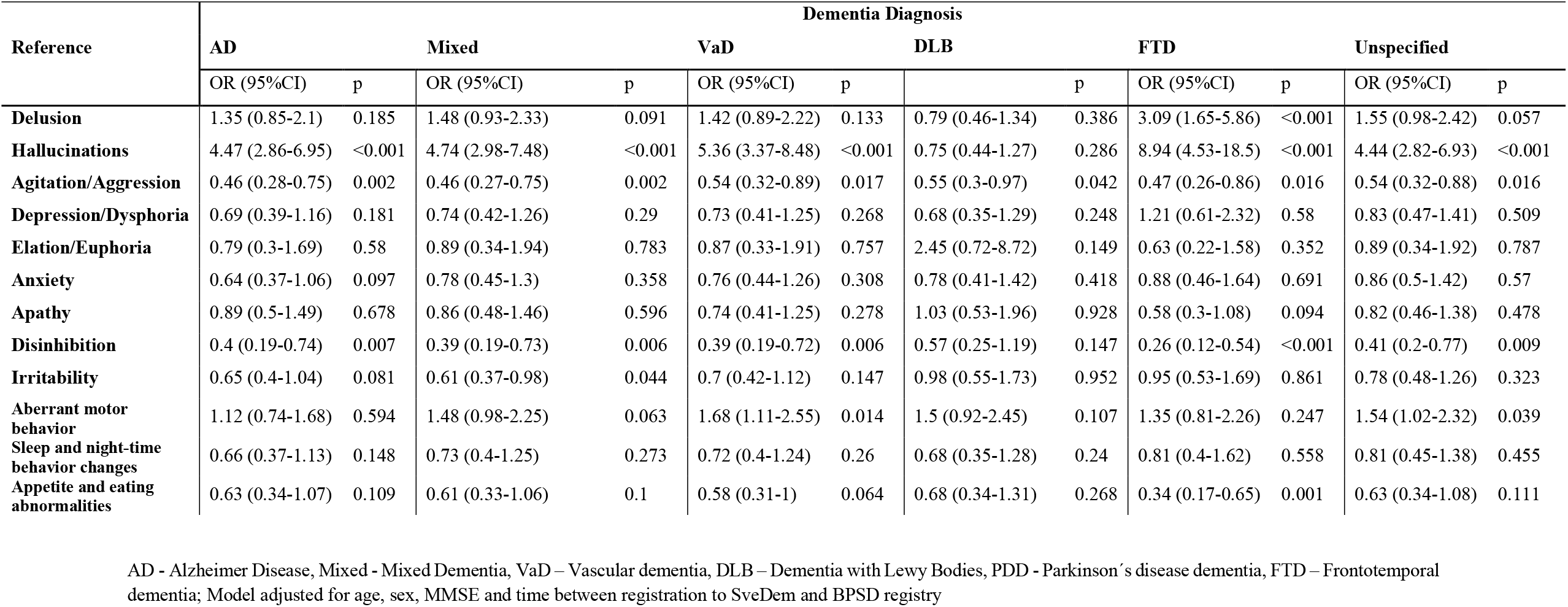
ODDS RATIOS (OR) AND 95% CONFIDENCE INTERVALS (CI) OF THE ASSOCIATION BETWEEN PDD AND BPSD IN PDD COMPARED TO ALL OTHER DIAGNOSES.

**Supplementary Table 2f.**
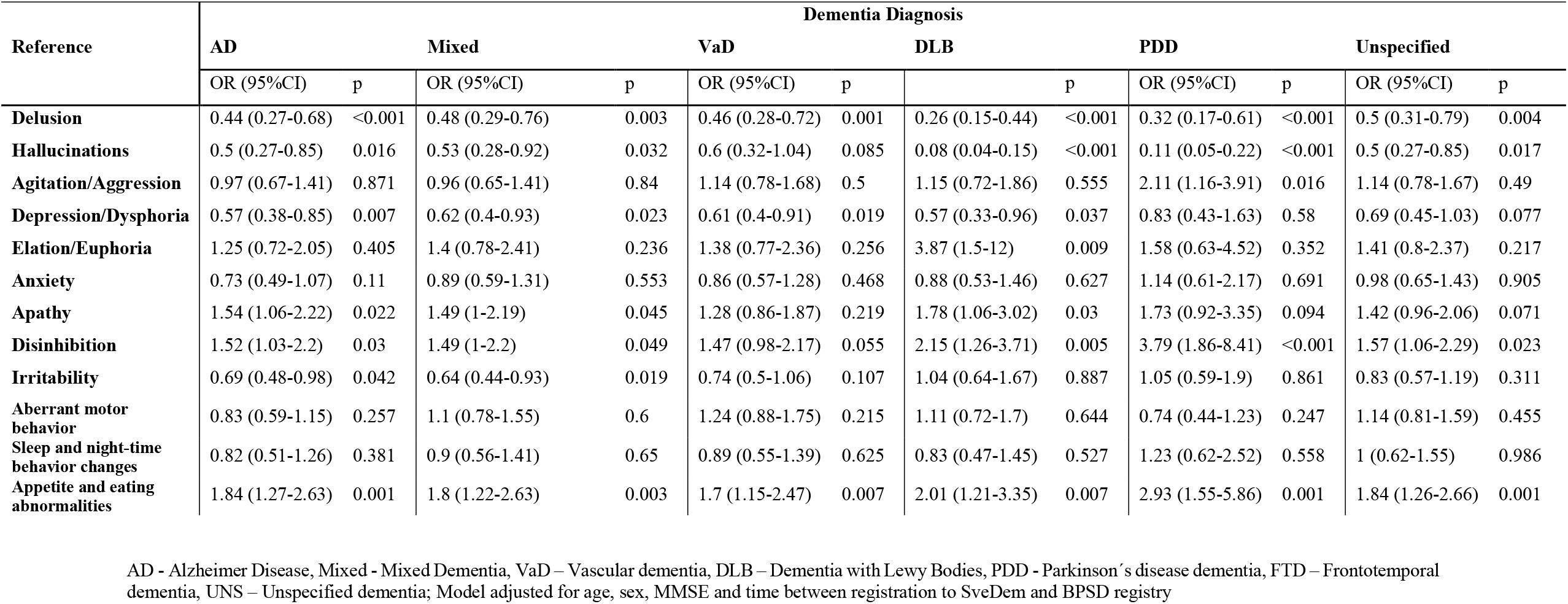
ODDS RATIOS (OR) AND 95% CONFIDENCE INTERVALS (CI) OF THE ASSOCIATION BETWEEN FTD AND BPSD IN FTD COMPARED TO ALL OTHER DIAGNOSES.

**Supplementary Table 2g.**
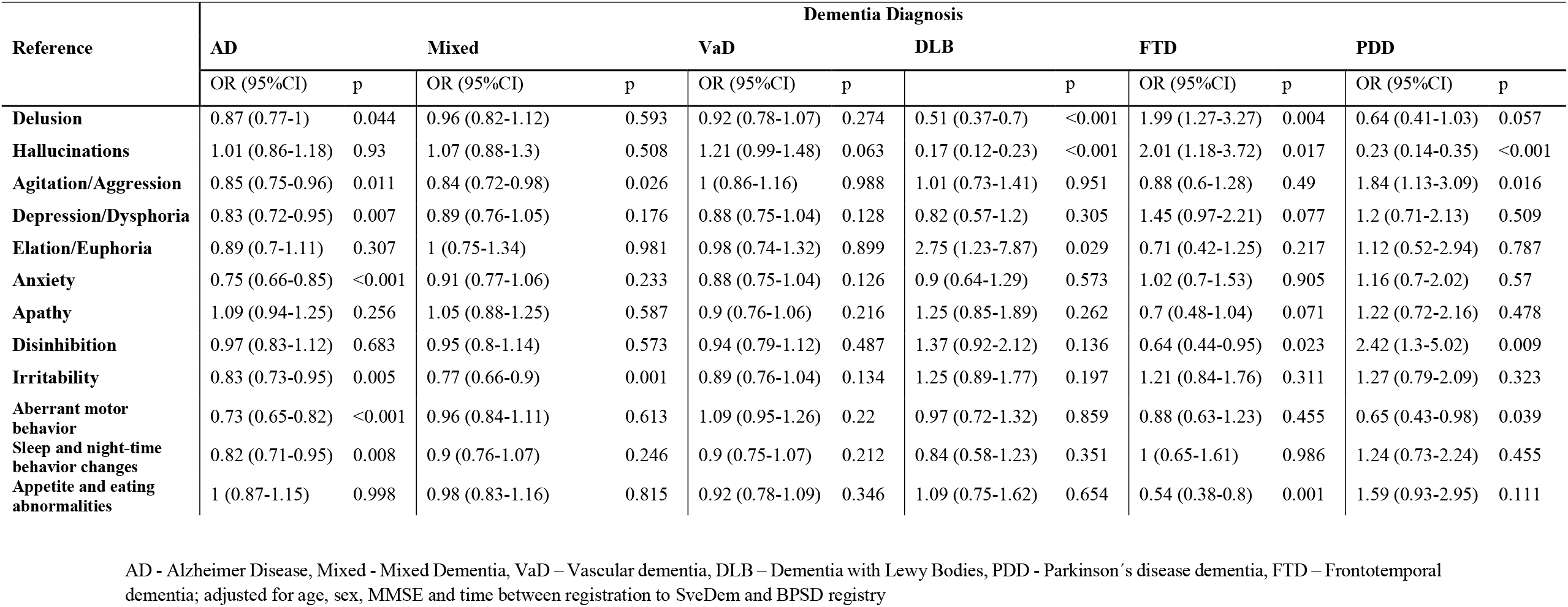
ODDS RATIOS (OR) AND 95% CONFIDENCE INTERVALS (CI) OF THE ASSOCIATION BETWEEN UNSPECIFIED DEMENTIA AND BPSD COMPARED TO ALL OTHER DIAGNOSES.

